# Understanding COVID-19 Vaccine Uptake and Hesitancy Among People With HIV in Freetown, Sierra Leone: A Cross-sectional Study

**DOI:** 10.1101/2023.05.11.23289882

**Authors:** Peterlyn E. Cummings, Sulaiman Lakoh, Sahr A. Yendewa, Samuel P.E. Massaquoi, Peter B. James, Foday Sahr, Gibrilla F. Deen, Pelema Gevao, George A. Yendewa

## Abstract

**Objectives:** People living with HIV (PWH) are at increased risk of COVID-19 related morbidity and mortality, yet less is known about COVID-19 vaccination uptake and hesitancy, especially in sub-Saharan Africa. We aimed to evaluate COVID-19 vaccine uptake and hesitancy among PWH in Sierra Leone.

**Methods:** We conducted a cross-sectional study in a convenience sample of PWH in routine care at Connaught Hospital in Freetown, Sierra Leone from April through June 2022. We collected sociodemographic and health-related data. We used the VAX Scale, a validated instrument to assess attitudes towards COVID-19 vaccination. From the responses, we constructed hesitancy (VAX) scores, with higher scores implying negative attitudes towards vaccination. We used generalized linear models to identify factors associated with vaccine hesitancy.

**Results:** A total of 490 PWH were enrolled (71.4% female, median age 38 years, median CD4 count 412 cells/mm^3^, 83.9% virologically suppressed). About 17.3% had received at least one dose of a COVID-19 vaccine. The mean VAX score was 43.14 ± 7.05, corresponding to 59.9% participants classified as vaccine hesitant. Preference for natural immunity (65.8%) and concerns about commercial profiteering (64.4%) were the commonest reasons for hesitancy, followed by mistrust of vaccine benefits (61.4%) and worries about future side effects (48.0%). In adjusted regression analysis, being Muslim (β = 2.563, p < 0.001) and residence in urban areas (β = 1.709, p = 0.010) were associated with greater vaccine hesitancy, while having tested ever for COVID-19 was associated with lesser vaccine hesitancy (β = -3.417, p = 0.027).

**Conclusion:** We observed a low COVID-19 vaccine uptake and high hesitancy among PWH in Sierra Leone. Our findings underscore the need to address vaccine hesitancy as a critical element of efforts to boost COVID-19 vaccine uptake among this population in Sierra Leone.

## INTRODUCTION

More than three years since the first cases were reported, the coronavirus disease 2019 (COVID-19) pandemic, caused by severe acute respiratory syndrome coronavirus 2 (SARS-CoV-2), continues to pose a significant global challenge, with millions of confirmed cases and documented deaths reported worldwide [1]. Unlike previous health crises of similar scale, the development of vaccines has occurred relatively early in the pandemic, leading to a significant shift in its trajectory. Current COVID-19 vaccines have proven to be effective in reducing virus transmission [2, 3], severity of illness [4], and COVID-19-associated mortality [5]. COVID-19 vaccination is now recommended for most individuals including people living with HIV (PWH), who are a priority population for vaccination. Compared with their non-HIV counterparts, PWH may be at increased susceptibility to severe illness and poor outcomes from SARS-CoV-2 infection, partly due to having a higher prevalence of premorbid risk factors including cardiovascular diseases, diabetes, and obesity [6, 7].

Despite ongoing efforts to promote COVID-19 vaccination, hesitancy to vaccinate persists among various populations, including PWH. Studies have provided insights into COVID-19 vaccination status and hesitancy among PWH. In a large global HIV cohort enrolled in the REPRIEVE study (n= 6952), Fulda et al [8] reported a COVID-19 vaccination rate of 55%, with large disparities in vaccination coverage rates noted between high-income countries in North America and Europe (71%) and low-income countries in Sub-Saharan Africa (18%). In a separate study assessing reasons for COVID-19 vaccine hesitancy among PWH (n=1030), Shrestha et al [9] found that up to 90% of survey respondents in the United States experienced some degree of COVID-19 vaccine hesitancy, with greater reluctance to vaccinate associated with being Black, holding conservative political viewpoints, and having concerns about vaccine safety and efficacy. However, the determinants of COVID-19 vaccine hesitancy among PWH in low-income settings are understudied, especially in sub-Saharan African countries where the global burden of HIV is highest.

With the emergence of more infectious variants of SARS-CoV-2, PWH in regions with a high burden of HIV may face continued risks of COVID-19-related morbidity and mortality, which may necessitate additional booster doses of COVID-19 vaccination. Understanding and addressing the factors contributing to vaccine hesitancy among PWH is essential to enhance vaccination rates in this population. In this study, we aimed to assess COVID-19 vaccination coverage and hesitancy in a cohort of PWH in Freetown, Sierra Leone.

## METHODS

### Study design, population, and setting

We conducted a cross-sectional study to assess COVID-19 vaccination coverage and vaccine hesitancy among PWH who received routine clinical care at the HIV Clinic at Connaught Hospital in Freetown, Sierra Leone from April through June 2022. The HIV Clinic is the largest HIV treatment center in Sierra Leone and has over 4000 PWH in active clinical follow-up. Connaught Hospital is affiliated with the College of Medicine and Allied Health Sciences of the University of Sierra Leone. The study inclusion criteria were (1) age ≥ 18 years, (2) documented evidence of HIV infection, and (3) willingness to give informed consent. The exclusion criteria were (1) age < 18 years, and (2) unwillingness or inability to give informed consent. Eligible patients were approached during routine clinic visits and informed of the purpose of the study. We used convenience sampling to enroll interested participants.

### Sample size calculation and justification

We estimated the minimum sample size n, according to Lwanga and Lemeshow [10], as follows:

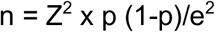

where Z = 1.96 at 95% confidence interval (CI), p = prevalence of COVID-19 vaccine hesitancy among PWH in Sierra Leone, and e is the error rate. Given the lack of studies among PWH from Sierra Leone, we used a COVID-19 vaccine hesitancy rate of 50% and an error rate of 5%, which yielded a sample size of 384. Factoring in a 10% non-response rate yielded a final minimum sample size of 422, which was sufficiently powered at 80% to detect associations between variables using a 2-tailed test.

### Survey instrument, procedures and measures

We collected baseline data from survey participants. Sociodemographic variables included age, sex, highest education attained, occupation, and religion. HIV-specific data included the most recent CD4 count (dichotomized as < 200 cells/mm3 vs ≥ 200 cells/mm3), HIV viral load (dichotomized as < 1000 copies/mL vs ≥ 1000 copies/mL), antiretroviral therapy (ART), and duration since HIV diagnosis. For COVID-19-related experiences, we collected data on SARS-CoV-2 testing history, COVID-19 vaccination history (number of doses received and type of vaccine), and vaccine-related adverse effects, with responses recorded as “yes” or “no”.

The instrument for assessing COVID-19 vaccine hesitancy was adapted from the 12-item Vaccination Attitudes Examination (VAX) scale originally developed by Martin and Petrie [11], which we have previously validated in Sierra Leone to assess COVID-19 vaccine among healthcare workers [12]. Briefly, the VAX Scale assesses attitudes towards vaccines across four domains, as follows: (1) mistrust of vaccine benefits, (2) worries about unforeseen future effects, (3) concerns about commercial profiteering, and (4) preference for natural immunity [11]. Items are rated on a six-point Likert scale with equidistant scores, as follows: 1 = strongly agree, 2 = agree, 3 = slightly agree, 4 = slightly disagree, 5= disagree and 6 = strongly disagree. The instrument was first piloted to the target study population (n=10) to ensure clarity of items. The pilot survey participants were not included in the final study.

We estimated the prevalence of COVID-19 vaccine hesitancy by summing participant responses to each item on the VAX scale. As items 4-12 were negatively worded, we reverse-scored responses to ensure that all items were keyed in a positive direction, with higher VAX scores indicating greater reluctance to vaccinate against COVID-19. The possible VAX scores ranged from 12 (positive attitude) to 72 (negative attitude). As previously described by us and others [12, 13], VAX scores ranging from 12-32 (i.e., 25th percentile) were categorized as low hesitancy, scores ranging from 33-52 (i.e., 50th percentile) were classified as moderate hesitancy, while scores > 52 (i.e., 75th percentiles) indicated high COVID-19 hesitancy. Vaccine hesitancy was defined as VAX score > mean (i.e., 50th percentile). Normality of the VAX score distribution was assessed using the Kolmogorov–Smirnov test, which assumed homogeneity of variances under the null hypothesis.

To assess the psychometric properties of the survey instrument, we estimated the internal consistency of the responses using mean inter-item reliability correlations and Cronbach’s alpha coefficients (α), with an overall α > 0.7 regarded as acceptable. We performed exploratory factor analysis using principal axis factoring with orthogonal (Varimax) rotation to assess the dimensional structure of the VAX scale.

### Statistical analysis

Statistical analyses were performed using the SPSS Version 29.0 (Armonk, NY, USA; IBM Corp). Categorical variables were reported as frequencies (percentages) and continuous variables as means (standard deviation) or medians (range or interquartile range, IQR). Generalized linear regression models were used to identify factors associated with COVID-19 vaccine hesitancy, represented by VAX scores. The covariates tested included sociodemographic and clinical data as described earlier and were included in multivariable model if significant in the univariate model. In all analysis, statistical significance was set at p < 0.05.

### Ethical approval

Ethical approval was obtained from the Sierra Leone Ethics and Scientific Review Committee (approval date 20 December 2021). Written informed consent was obtained from each participant before enrolment into the study.

## RESULTS

### Characteristics of Participants

A total of 490 PWH participated in the study (Table 1), of which 71.4% (350/490) were female. The median age was 38 years (IQR 32-49), and the majority were single (60.6%, 297/490), employed in the informal sector (69.4%, 340/490), and Muslim (69.2%, 339/490). Most (80%, 392/490) had attained primary education or higher. The median CD4 count was 412 cells/mm3 (IQR 256-508). Most (83.9%, 411/490) were virologically suppressed (< 1000 copies/mL) and on dolutegravir-based ART (57.8%, 283/490).

**Table 1.** Sociodemographic and health characteristics of participants (N=490)

| Characteristics | N (%) |
| --- | --- |
| **Gender** |  |
| Male | 140 (28.6) |
| Female | 350 (71.4) |
| **Age, years** |  |
| Median (IQR) | 38 (32-49) |
| <25 | 16 (3.3) |
| 25-34 | 144 (29.4) |
| 35-44 | 167 (34.1) |
| 45-54 | 99 (20.2) |
| ≥55 | 64 (13.1) |
| **Relationship status** |  |
| Single | 297 (60.6) |
| Married | 155 (31.6) |
| Widowed/separated | 38 (7.8) |
| **Highest education attained** |  |
| None | 98 (20.0) |
| Primary | 111 (22.7) |
| Secondary | 225 (45.9) |
| Tertiary | 56 (11.4) |
| **Occupation** |  |
| Unemployed | 109 (22.2) |
| Informal | 340 (69.4) |
| Formal | 41 (8.4) |
| **Religion** |  |
| Christian | 151 (30.8) |
| Muslim | 339 (69.2) |
| **Time living with HIV, years** |  |
| Median (IQR) | 5 (3-11) |
| ≤ 2 | 69 (14.1) |
| 3-5 | 180 (36.7) |
| 6-10 | 109 (22.2) |
| ≥ 11 | 132 (26.9) |
| **Current CD4 count, cells/mm<sup>3</sup>** |  |
| Median | 412 (256-508) |
| <200 | 79 (16.1) |
| ≥200 | 405 (82.7) |
| **Viral load, copies/mL** |  |
| <1000 | 411 (83.9) |
| ≥1000 | 79 (16.1) |
| **Current ART** |  |
| Dolutegravir-based | 283 (57.8) |
| Efavirenz-based | 146 (29.8) |
| Lopinavir-based | 61 (12.4) |
| **History of chronic illness** |  |
| Yes | 24 (4.9) |
| No | 466 (95.1) |

### COVID-19 Vaccine Coverage and Experiences

As shown in Table 1, 17.3% (85/490) of participants received a COVID-19 vaccine. Of these, 63.5% (54/85) received 1 dose of a COVID-19 vaccine, while 36.5% (31/85) had received a complete series of vaccination (i.e., 2 doses). Furthermore, 38.8% (33/85) had the Janssen (Johnson & Johnson) vaccine, 32.9% (28/85) received the AstraZeneca vaccine, and 28.2% (24/85) received the Sinopharm vaccine. About 44.6% (37/85) reported experiencing at least one vaccine-related adverse effect, as follows (not mutually exclusive): pain at the injection site (38.8%, 33/85), body aches (9.4%, 8/85) and fever (3.5%, 3/85). Overall, only 5.7% (28/490) had ever been tested for SARS-CoV-2 infection, of which no positive cases were detected.

### Prevalence of COVID-19 Vaccine Hesitancy

Participants’ responses to items on the VAX scale demonstrated excellent internal consistency (Cronbach’s α = 0.94 overall, range 0.88-0.94 across domains) (Table 2). Similarly, the mean inter-item correlation was high (r = 0.645 overall, range 0.745-0.892 across domains). The VAX scores ranged from 27-60 and the Kolmogorov–Smirnov test indicated that the VAX scores were not normally distributed (K-S = 0.12, p < 0.001). Exploratory factor analysis with principal axis factoring using Varimax rotation confirmed the 4-factor solution of the original VAX scale by Martin and Petrie [11].

**Table 2.** Summary of COVID-19 attitude statements and vaccine hesitancy levels (in percentages, %)

| COVID-19 Vaccine Attitude Statements | Vaccine Hesitancy Level |  |  | Mean Inter-Item Correlation | Domain Cronbach's Alpha |
| --- | --- | --- | --- | --- | --- |
|  | Low | Moderate | High |  |  |
| **Mistrust of vaccine benefits** |  |  |  |  |  |
| I feel that the COVID-19 vaccine is very safe | 37.9 | 12.0 | 50.1 |  |  |
| I can rely on the COVID-19 vaccine to prevent serious infection with COVID-19 | 38.1 | 11.8 | 49.8 | 0.892 | 0.94 |
| I feel fully protected from COVID-19 infection in the future after getting the COVID-19 vaccine | 31.6 | 16.7 | 51.7 |  |  |
| **Worries about unforeseen future effects** |  |  |  |  |  |
| Although the COVID-19 vaccine appears to be safe, there may be problems with the vaccine that we have not yet discovered (r) | 56.3 | 17.0 | 26.7 |  |  |
| The COVID-19 vaccine can cause unforeseen problems in the future (r) | 54.7 | 17.8 | 27.1 | 0.823 | 0.90 |
| I worry about the unknown future effects of the COVID-19 vaccine (r) | 68.8 | 6.7 | 24.5 |  |  |
| **Concerns about commercial profiteering** |  |  |  |  |  |
| COVID-19 vaccine will make a lot of money for pharmaceutical companies but will not bring much benefit to common people (r) | 37.7 | 13.9 | 48.4 |  |  |
| Authorities promote the COVID-19 vaccine for financial gain, not for people's health (r) | 32.0 | 12.6 | 55.4 | 0.745 | 0.88 |
| COVID-19 vaccination programs are a fraud (r) | 29.4 | 6.8 | 63.8 |  |  |
| **Preference for natural immunity** |  |  |  |  |  |
| Natural immunity will last longer than immunity from the COVID-19 vaccine (r) | 31.8 | 18.2 | 50.0 |  |  |
| Natural exposure to the virus gives the safest protection against COVID-19 (r) | 24.3 | 29.8 | 45.9 | 0.846 | 0.93 |
| Being exposed to COVID-19 naturally is safer for the immune system than being exposed through vaccination (r) | 23.3 | 19.2 | 57.5 |  |  |

Abbreviations: (r), reverse-scored in a positive direction on a 6-point Likert scale ranging from 1 (strongly disagree) to 6 (strongly agree) and reclassified hesitancy as low (1 and 2), moderate (3 and 4) and high (5 and 6)

The overall mean VAX score was 43.14 ± 7.05, corresponding to 59.9% of participants expressing COVID-19 hesitancy (Table 3). Across domains, preference for natural immunity (65.8%, mean score 11.84 ± 3.89) and concerns for commercial profiteering (64.4%, mean score 11.62 ± 4.11) were the most common reasons for vaccine hesitancy, followed by mistrust of vaccine benefits (61.4%, mean score 11.06 ± 4.59) and worries about unforeseen future effects (48.0%, mean score 8.62 ± 4.05). Furthermore, 6.7%, 91.0%, and 2.3% of participants were classified into the low-, mild-to-moderate- and high-level vaccine hesitancy categories, respectively.

**Table 3.** Domain and Overall Vaccine Hesitancy Scores

| Variables | Expected Range of VAX Score | VAX Score or N | % Participants With VAX Score |
| --- | --- | --- | --- |
| **Overall hesitancy** |  |  |  |
| Mean (SD) | 12-72 | 43.14 ± 7.05 | 59.9 |
| **Hesitancy domains** |  |  |  |
| Mistrust of vaccine benefits | 3-18 | 11.06 ± 4.59 | 61.4 |
| Worries about unforeseen future effects | 3-18 | 8.62 ± 4.05 | 48.0 |
| Concerns about commercial profiteering | 3-18 | 11.62 ± 4.11 | 64.4 |
| Preference for natural immunity | 3-18 | 11.84 ± 3.89 | 65.8 |
| **Categories of hesitancy** |  |  |  |
| Low | 12-32 | 33 | 6.7 |
| Moderate | 33-52 | 446 | 91.0 |
| High | >52 | 11 | 2.3 |

Abbreviations: N, sample size; Min, minimum; Max, maximum; SD, standard deviation; VAX Score, vaccine hesitancy score

### Factors Associated With COVID-19 Vaccine Hesitancy

In the univariate and multivariable linear regression analyses (Table 4), greater COVID-19 vaccine hesitancy was associated with being Muslim (β = 2.563, p < 0.001) and residence in urban areas (β = 1.709, p = 0.010), while having ever tested for COVID-19 was associated with less vaccine hesitancy (β = -3.417, p = 0.027).

**Table 4.** Univariate and multivariable linear regression correlates of COVID-19 vaccine hesitancy

| Variables | Univariate |  |  | Multivariable |  |  |
| --- | --- | --- | --- | --- | --- | --- |
| | $\beta$ | S.E. | *p*-Value | $\beta$ | S.E. | *p*-Value |
| **Sociodemographic information** |  |  |  |  |  |  |
| Sex: male | -0.896 | 0.702 | 0.202 |  |  |  |
| Age (years) | 0.040 | 0.030 | 0.179 |  |  |  |
| Relationship status: single | -0.010 | 0.648 | 0.988 |  |  |  |
| Education: none | 0.661 | 0.791 | 0.403 |  |  |  |
| Religion: Muslim | 2.511 | 0.676 | **<0.001** | 2.563 | 0.674 | **<0.001** |
| Unemployed | 0.552 | 0.761 | 0.469 |  |  |  |
| Residence: urban | 1.476 | 0.671 | **0.028** | 1.709 | 0.661 | **0.010** |
| **HIV-related factors** |  |  |  |  |  |  |
| Time living with HIV | 0.119 | 0.074 | 0.107 |  |  |  |
| CD4 count | 0.001 | 0.001 | 0.293 |  |  |  |
| Viral load: suppressed | 0.474 | 0.861 | 0.582 |  |  |  |
| ART class: dolutegravir-based | 1.179 | 0.640 | 0.065 |  |  |  |
| **COVID-19 and other health information** |  |  |  |  |  |  |
| Ever been tested for COVID-19 | -3.417 | 1.355 | **0.012** | -2.943 | 1.334 | **0.027** |
| History of chronic illness | 0.356 | 1.467 | 0.808 |  |  |  |

$\beta$ , estimate; S.E., standard error

## DISCUSSION

Despite being vulnerable to poor outcomes from COVID-19, there are limited studies on COVID-19 vaccine uptake and hesitancy among PWH in sub-Saharan Africa. To effectively address this issue in the region, it is crucial to measure and understand the extent and underlying reasons for this phenomenon. Our study revealed a concerningly low COVID-19 vaccine coverage (17.3%) and a high level of vaccine hesitancy (59.9%) among PWH in an urban setting in Sierra Leone. Similar findings were reported in a study from South Africa (n=213) by Govere-Hwenje et al [14], where 57% of PWH indicated willingness to accept COVID-19 vaccination, while 21% were unwilling to vaccinate and 20% were unsure. Another study (n=660) by Sulaiman et al [15] observed a 58% COVID-19 vaccine hesitancy rate among PWH across six hospital systems in Nigeria. In contrast, Muhindo et al [16] found a high COVID-19 vaccination coverage (69.6%) and high confidence in the vaccine in a Ugandan HIV cohort. Overall, however, these studies highlight the persistently high levels of COVID-19 vaccine hesitancy among PWH, despite concerted efforts to increase vaccine uptake in this population.

The most common reasons for vaccine hesitancy among our study participants were a preference for natural immunity (65.4%) and concerns about commercial profiteering/mistrust of pharmaceutical industries (64.4%). Notably, fewer participants expressed concerns about potential vaccine side effects (48.0%), in contrast with our previous study among healthcare workers in Sierra Leone which was conducted during the same timeframe as the present study (n=592), where 76% expressed worries about vaccine side effects [12]. We hypothesize that HIV-positive status may introduce additional concerns including uncertainty about vaccine safety and efficacy and the possibility of vaccine interactions with HIV medications, which may affect the immune system [17, 18]. Additionally, PWH may have negative experiences, including stigmatization, while navigating the healthcare system, which can contribute to mistrust of the system and erosion of confidence in health authorities [19, 20]. The combined influence of these intersectional factors, along with misinformation and conspiracy theories, may contribute to vaccine hesitancy among PWH. Interestingly, however, contrary to other studies [9, 21], vaccine hesitancy was not predicted by HIV-specific factors such as CD4 count or viral load suppression.

Urban residence was associated with higher COVID-19 vaccine hesitancy. However, a comprehensive study of the general population in five West African countries (Burkina Faso, Guinea, Mali, Senegal, and Sierra Leone) with a large sample size (n=4198) revealed no significant association between vaccine acceptance and urban versus rural residence [22]. These contrasting findings may be attributed to various social, cultural, and economic factors that influence vaccine attitudes and behaviors in different settings. Urban areas, with their diverse populations comprising individuals with varying levels of education, income, and healthcare access, may have more complex vaccine decision-making processes [22]. Additionally, urban areas may be more susceptible to misinformation and conspiracy theories about vaccine safety and efficacy, contributing to vaccine hesitancy [23]. Further research is needed to explore the complex relationships between area of residence, vaccine hesitancy, and other socio-demographic factors.

Furthermore, it was noteworthy that being Muslim was a predictor of COVID-19 vaccine hesitancy in our study. This finding is in line with previous reports from and outside sub-Saharan Africa which have observed higher levels of hesitancy among Muslim populations to vaccination against COVID-19 and other diseases [17, 24, 25]. However, vaccine hesitancy has been found among Christians and people of all religious backgrounds, suggesting that the relationship between religious identity, religiosity, and vaccine hesitancy is not universal and may be influenced by cultural, socioeconomic, and geographic contexts [26-28]. Thus, efforts to address vaccine hesitancy among populations should focus on providing accurate information, addressing misconceptions, and building trust in the safety and efficacy of vaccines within these communities. Collaborative initiatives with trusted religious leaders, community organizations, and healthcare providers who understand the cultural and religious nuances can be effective in addressing vaccine hesitancy among populations with diverse religious and cultural backgrounds [24, 29].

Another important finding was that having ever tested for COVID-19 was associated with lower vaccine hesitancy. This may be explained by theories of health behavior, which posit that positive health behavior may be elicited by first-hand experiences with illness [30-32]. For example, a personal experience of testing positive for COVID-19 or witnessing loved ones’ illness may enhance an individual’s perception of the severity of the infection, resulting in a heightened sense of urgency to protect oneself and others through vaccination [33]. Additionally, testing efforts often provide accurate information about the virus and vaccines, which can address misinformation and misconceptions that contribute to vaccine hesitancy [34]. Moreover, the emphasis on collective responsibility and community health in testing efforts may foster a sense of social obligation to take preventive measures, including vaccination [34, 35].

Our study was characterized by strengths and limitations. Firstly, our survey utilized convenience sampling, which may have led to an underestimation of the true prevalence of COVID-19 vaccine hesitancy. Secondly, our study was limited to PWH in an urban setting and may not be representative of attitudes towards COVID-19 vaccination nationally. Thirdly, our investigation of barriers to vaccine uptake was not comprehensive and could be better explored using qualitative or mixed-methods study designs which are better suited to explore causal links. Nevertheless, our study adds to our understanding of COVID-19 vaccination uptake and hesitancy among PWH in Sierra Leone and can help guide interventions aimed at improving COVID-19 vaccine acceptance in this population.

## CONCLUSION

In summary, we observed a low high prevalence of COVID-19 vaccination and high levels of vaccine hesitancy among PWH in Freetown Sierra Leone. The most frequent reasons for COVID-19 vaccine hesitancy were preference for natural immunity and concerns about commercial profiteering, followed by mistrust of vaccine benefits and worries about future side effects. Being Muslim and urban residence were associated with greater vaccine hesitancy, while having ever tested for COVID-19 was associated with lesser vaccine hesitancy. Given that PWH remain a venerable population to poor COVID-19 outcomes, our findings may help inform strategies aimed at increasing COVID-19 vaccine uptake in this setting.

## Data Availability

All data produced in the present study are available upon reasonable request to the authors

## AUTHOR CONTRIBUTIONS

GAY, PEC, SL and PG conceptualized and designed the study. SAY, SPEM, and PBJ contributed to study concept and design. PEC collected the data. GAY conducted the statistical analysis. FS and GFD contributed important intellectual content. All authors contributed to the interpretation of data. GAY wrote the initial manuscript draft. All authors critically revised and approved of the final version. GAY is acting as the guarantor of this manuscript.

## ACKLOWLEDGEMENTS

We wish to acknowledge people living with HIV and clinic staff at Connaught Hospital in Freetown, Sierra Leone, without whose help this study would not have been successful.

## DATA AVAIL ABILITY STATEMENT

The data presented in this study are available on request from the corresponding author upon reasonable request.

## ETHICAL APPROVAL

Ethical approval was obtained from the Sierra Leone Ethics and Scientific Review Committee (approval date 20 December 2021). Written informed consent was obtained from participants prior to enrolment into the study.

## REFERENCES

1. WHO 2023. Coronavirus disease (COVID-19) Weekly Epidemiological Updates and Monthly Operational Updates. Available online: https://www.who.int/emergencies/diseases/novel-coronavirus-2019/situation-reports (accessed 25 April 2023).

2. Singanayagam A, Hakki S, Dunning J, Madon KJ, Crone MA, Koycheva A, Derqui-Fernandez N, Barnett JL, Whitfield MG, Varro R, Charlett A, Kundu R, Fenn J, Cutajar J, Quinn V, Conibear E, Barclay W, Freemont PS, Taylor GP, Ahmad S, Zambon M, Ferguson NM, Lalvani A; ATACCC Study Investigators. Community transmission and viral load kinetics of the SARS-CoV-2 delta (B.1.617.2) variant in vaccinated and unvaccinated individuals in the UK: a prospective, longitudinal, cohort study [published correction appears in Lancet Infect Dis. 2021;21(12):e363]. Lancet Infect Dis. 2022;22(2):183–195.

3. Salo J, Hägg M, Kortelainen M, Leino T, Saxell T, Siikanen M, Sääksvuori L. The indirect effect of mRNA-based COVID-19 vaccination on healthcare workers’ unvaccinated household members. Nat Commun. 2022;13(1):1162.

4. Tenforde MW, Self WH, Adams K, Gaglani M, Ginde AA, McNeal T, Ghamande S, Douin DJ, Talbot HK, Casey JD, Mohr NM, Zepeski A, Shapiro NI, et al. Association Between mRNA Vaccination and COVID-19 Hospitalization and Disease Severity. JAMA. 2021;326(20):2043–2054.

5. Kelly JD, Leonard S, Hoggatt KJ, Boscardin WJ, Lum EN, Moss-Vazquez TA, Andino R, Wong JK, Byers A, Bravata DM, Tien PC, Keyhani S. Incidence of Severe COVID-19 Illness Following Vaccination and Booster With BNT162b2, mRNA-1273, and Ad26.COV2.S Vaccines. JAMA. 2022;328(14):1427–1437.

6. Yendewa GA, Perez JA, Schlick K, Tribout H, McComsey GA. Clinical Features and Outcomes of Coronavirus Disease 2019 Among People With Human Immunodeficiency Virus in the United States: A Multicenter Study From a Large Global Health Research Network (TriNetX). Open Forum Infect Dis. 2021;8(7):ofab272.

7. Bhaskaran K, Rentsch CT, MacKenna B, Schultze A, Mehrkar A, Bates CJ, Eggo RM, Morton CE, Bacon SCJ, Inglesby P, Douglas IJ, Walker AJ, McDonald HI, Cockburn J, Williamson EJ, Evans D, Forbes HJ, Curtis HJ, Hulme WJ, Parry J, Hester F, Harper S, Evans SJW, Smeeth L, Goldacre B. HIV infection and COVID-19 death: a population-based cohort analysis of UK primary care data and linked national death registrations within the OpenSAFELY platform. Lancet HIV. 2021;8(1):e24–e32.

8. Fulda ES, Fitch KV, Overton ET, Zanni MV, Aberg JA, Currier JS, Lu MT, Malvestutto C, Fichtenbaum CJ, Martinez E, Umbleja T, Douglas PS, Ribaudo HJ, Grinspoon SK. COVID-19 Vaccination Rates in a Global HIV Cohort. J Infect Dis. 2022;225(4):603–607.

9. Shrestha R, Meyer JP, Shenoi S, Khati A, Altice FL, Mistler C, Aoun-Barakat L, Virata M, Olivares M, Wickersham JA. COVID-19 Vaccine Hesitancy and Associated Factors among People with HIV in the United States: Findings from a National Survey. Vaccines (Basel). 2022;10(3):424.

10. Lwanga SK, Lemeshow S, World Health Organization (1991). Sample size determination in health studies: a practical manual. Available online: https://apps.who.int/iris/handle/10665/40062 (cited 24 April 2023).

11. Martin LR, Petrie KJ. Understanding the Dimensions of Anti-Vaccination Attitudes: the Vaccination Attitudes Examination (VAX) Scale. Ann Behav Med. 2017; 51(5):652–660.

12. Yendewa SA, Ghazzawi M, James PB, Smith M, Massaquoi SP, Babawo LS, Deen GF, Russell JBW, Samai M, Sahr F, Lakoh S, Salata RA, Yendewa GA. COVID-19 Vaccine Hesitancy among Healthcare Workers and Trainees in Freetown, Sierra Leone: A Cross-Sectional Study. Vaccines (Basel). 2022;10(5):757.

13. Oke OO, Akinboro AO, Olanrewaju FO, Oke OA, Omololu AS. Assessment of HIV-related stigma and determinants among people living with HIV/AIDS in Abeokuta, Nigeria: A cross-sectional study. SAGE Open Med. 2019;7:2050312119869109.

14. Govere-Hwenje S, Jarolimova J, Yan J, Khumalo A, Zondi G, Ngcobo M, Wara NJ, Zionts D, Bogart LM, Parker RA, Bassett IV. Willingness to accept COVID-19 vaccination among people living with HIV in a high HIV prevalence community. BMC Public Health. 2022;22(1):1239.

15. Kabir Sulaiman S, Sale Musa M, Isma’il Tsiga-Ahmed F, Muhammad Dayyab F, Kabir Sulaiman A, Dabo B, Idris Ahmad S, Abubakar Haruna S, Abdurrahman Zubair A, Hussein A, Usman S, Usman Wada J, Yekeen Ayodele A, Wulgo Ali M, Tijjani Makama B, Tijjani Bako A; SQuAD-HIV collaborators. COVID-19 vaccine hesitancy among people living with HIV in a low-resource setting: A multi-center study of prevalence, correlates and reasons. Vaccine. 2023;41(15):2476–2484.

16. Muhindo R, Okoboi S, Kiragga A, King R, Arinaitwe WJ, Castelnuovo B. COVID-19 vaccine acceptability, and uptake among people living with HIV in Uganda. PLoS One. 2022;17(12):e0278692.

17. Iliyasu Z, Kwaku AA, Umar AA, Tsiga-Ahmed F, Nass NS, Abdullahi HM, Amole TG, Salihu HM, Aliyu MH. Predictors of COVID-19 Vaccine Acceptability among Patients Living with HIV in Northern Nigeria: A Mixed Methods Study. Curr HIV Res. 2022;20(1):82–90.

18. Mohamed R, White TM, Lazarus JV, Salem A, Kaki R, Marrakchi W, Kheir SGM, Amer I, Ahmed FM, Khayat MA, Al-Abdullah N, Ali B, Sultan R, Alamri B, Abdulmajid A, Kooli I, Chakroun M, Madani TA, Esmat G, Cordie A. COVID-19 vaccine acceptance and associated factors among people living with HIV in the Middle East and North Africa region. South Afr J HIV Med. 2022;23(1):1391.

19. Turan JM, Elafros MA, Logie CH, Banik S, Turan B, Crockett KB, Pescosolido B, Murray SM. Challenges and opportunities in examining and addressing intersectional stigma and health. BMC Med. 2019;17(1):7.

20. Maragh-Bass AC, Aimone EV, Aikhuele EO, Macqueen K. Exploring intersectional stigma and COVID-19 impact on human immunodeficiency virus service provision for African Americans in a Southern city. J Clin Nurs. 2022;10.1111/jocn.16539.

21. Balaji JN, Prakash S, Joshi A, Surapaneni KM. A Scoping Review on COVID-19 Vaccine Hesitancy among the Lesbian, Gay, Bisexual, Transgender, Queer, Intersex and Asexual (LGBTQIA+) Community and Factors Fostering Its Refusal. Healthcare (Basel). 2023;11(2):245.

22. Faye SLB, Krumkamp R, Doumbia S, Tounkara M, Strauss R, Ouedraogo HG, Sagna T, Barry AM, Mbawah AK, Doumbia CO, Diouf S, Cisse K, Harding M, Donven P, May J, Puradiredja DI, Fusco D; ACHES consortium. Factors influencing hesitancy towards adult and child COVID-19 vaccines in rural and urban West Africa: a cross-sectional study. BMJ Open. 2022;12(4):e059138.

23. Lv X, Zhao C, Song B, Huang H, Song S, Long H, Liu W, Du M, Liu M, Liu J. COVID-19 vaccination in people living with HIV and AIDS (PLWHA) in China: A cross-sectional study. Hum Vaccin Immunother. 2023;19(1):2151798.

24. Alsuwaidi AR, Hammad HAA, Elbarazi I, Sheek-Hussein M. Vaccine hesitancy within the Muslim community: Islamic faith and public health perspectives. Hum Vaccin Immunother. 2023;19(1):2190716.

25. Wonodi C, Obi-Jeff C, Adewumi F, Keluo-Udeke SC, Gur-Arie R, Krubiner C, Jaffe EF, Bamiduro T, Karron R, Faden R. Conspiracy theories and misinformation about COVID-19 in Nigeria: Implications for vaccine demand generation communications. Vaccine. 2022;40(13):2114–21.

26. Forster AS, Rockliffe L, Chorley AJ, Marlow LAV, Bedford H, Smith SG, Waller J. Ethnicity-specific factors influencing childhood immunisation decisions among Black and Asian Minority Ethnic groups in the UK: a systematic review of qualitative research. J Epidemiol Community Health. 2017;71(6):544–549.

27. Perry SL, Whitehead AL, Grubbs JB. Culture wars and COVID-19 conduct: Christian nationalism, religiosity, and Americans’ behavior during the coronavirus pandemic. Journal for the scientific study of religion. 2020;59(3):405–16.

28. Corcoran KE, Scheitle CP, DiGregorio BD. Christian nationalism and COVID-19 vaccine hesitancy and uptake. Vaccine. 2021;39(45):6614–21.

29. Olagoke AA, Olagoke OO, Hughes AM. Intention to Vaccinate Against the Novel 2019 Coronavirus Disease: The Role of Health Locus of Control and Religiosity. J Relig Health. 2021;60(1):65–80.

30. Noar SM, Zimmerman RS. Health Behavior Theory and cumulative knowledge regarding health behaviors: are we moving in the right direction? Health Educ Res. 2005;20(3):275–90.

31. Janz NK, Becker MH. The health belief model - a decade later. Health Ed Quart. 1984;11(1):1–47.

32. Rejeski WJ, Fanning J. Models and theories of health behavior and clinical interventions in aging: a contemporary, integrative approach. Clin Interv Aging. 2019;14:1007–1019.

33. Wickersham JA, Meyer JP, Shenoi S, Altice FL, Barakat LA, Virata M, Olivares M, Maviglia F, Khati A, Shrestha R. Willingness to Be Vaccinated Against COVID-19 Among People With HIV in the United States: Results From a National Survey. Front Med (Lausanne). 2022;9:886936.

34. Kricorian K, Civen R, Equils O. COVID-19 vaccine hesitancy: misinformation and perceptions of vaccine safety. Hum Vaccin Immunother. 2022;18(1):1950504.

35. Sergi CM, Leung AKC. Vaccination: a question of social responsibility. J Prev Med Hyg. 2021;62(1):E46–E47.

